# Modelling the impact of fexinidazole use on human African trypanosomiasis transmission in the Democratic Republic of Congo

**DOI:** 10.1101/2021.06.09.21258601

**Authors:** Aatreyee M. Das, Nakul Chitnis, Christian Burri, Daniel H. Paris, Swati Patel, Simon E.F. Spencer, Erick M. Miaka, M. Soledad Castaño

## Abstract

Gambiense human African trypanosomiasis is a deadly disease that has been declining in incidence since the start of the Century, primarily due to increased screening, diagnosis and treatment of infected people. The main treatment regimen currently in use requires a lumbar puncture as part of the diagnostic process to determine disease stage and hospital admission for drug administration. Fexinidazole is a new oral treatment for stage 1 and non-severe stage 2 human African trypanosomiasis. The World Health Organization has recently incorporated fexinidazole into its treatment guidelines for human African trypanosomiasis. The treatment does not require hospital admission or a lumbar puncture for all patients, which is likely to ease access for patients; however, it does require concomitant food intake, which is likely to reduce adherence. Here, we use a mathematical model calibrated to case and screening data from Mushie territory, in the Democratic Republic of the Congo, to explore the potential negative impact of poor compliance to an oral treatment, and potential gains to be made from increases in the rate at which patients seek treatment. We find that reductions in compliance in treatment of stage 1 cases are projected to result in the largest increase in further transmission of the disease, with failing to cure stage 2 cases also posing a smaller concern. Reductions in compliance may be offset by increases in the rate at which cases are passively detected. Efforts should therefore be made to ensure good adherence for stage 1 patients to treatment with fexinidazole and to improve access to passive care.

## 1 Introduction

Human African trypanosomiasis (HAT) is a vector-borne neglected tropical disease mainly affecting people in rural settings in sub-Saharan Africa. Two subspecies of *Trypanosoma brucei, T. b. gambiense* and *T. b. rhodesiense*, cause the slower and faster progressing forms of the disease, respectively. The *gambiense* form of the disease (gHAT) accounts for ∼ 98% of reported cases, with its greatest burden being in the Democratic Republic of the Congo (DRC) [1]. Nonetheless, the burden of disease has reduced significantly since the turn of the century, with the global number of cases reported to the World Health Organization (WHO) falling from 26,872 in 2001 to 876 in 2019 [2]. WHO has set the goal of interrupting transmission of gHAT by 2030 [3]. Actively screening at-risk populations for cases has formed the main control measure for gHAT.

Disease progression occurs in two stages: the first stage is the haemolymphatic stage, consisting of milder symptoms such as headaches and fever; the second stage is the meningoencephalitic stage, where the parasites cross the blood-brain barrier, leading to neuro-psychiatric disorders and eventual death if left untreated. Disease staging is required to define treatment, and is determined via examination of the cerebrospinal fluid obtained through a lumbar puncture. The recommended treatment for gHAT that is currently in use consists of daily intramuscular injection of pentamidine for seven days for stage 1 of the disease, and oral nifurtimox and intravenous eflornithine combination therapy (NECT) over ten days for stage 2, both requiring patient hospitalisation [4, 5].

Fexinidazole is a 10-day oral treatment for both stages of the disease. In November 2018, fexinidazole received a positive opinion by the European Medicines Agency for the treatment of both the first stage and second stage of gHAT in adults and children aged 6 years and older with a body weight of 20 kg or more. In December 2018, marketing authorisation was granted within the DRC. Recently, fexinidazole has been included in the WHO guidelines for the treatment of gHAT [4]. This new treatment presents significant advantages over the current treatment in terms of easier administration, a less unpleasant experience for patients and removing the need for a lumbar puncture in less severe cases. Although fexinidazole is an excellent drug for stage 1 and early stage 2 of the disease, a higher treatment failure rate was observed for late-stage 2 patients, where there is substantial parasite presence in the central nervous system. In these cases, treatment with NECT is recommended. Thus, patients only need a lumbar puncture if a clinical assessment suggests there is a chance of severe stage 2 HAT. As fexinidazole requires treatment for 10 consecutive days and food intake prior to drug administration to ensure efficacy, there is a concern that compliance may be lower than in the current treatment, which is parenterally administered by healthcare professionals. Finally, it is possible that, due to the easier logistics from a health facility perspective and less unpleasant treatment from a patient perspective, the rate of passive detection, i.e. detection at health center level and not by screening via a dedicated mobile team, may increase with the introduction of fexinidazole. This is in line with previous studies of patient preferences for anticancer treatments, which found a strong preference for oral as compared to parenterally administered chemotherapy [6, 7, 8].

In this study, we use a previously described and calibrated mathematical model for gHAT to explore how a reduced compliance to fexinidazole, as compared to the current treatment, may impact gHAT transmission. Furthermore, we consider the impact of potential increases in passive detection on mitigating the impact of non-compliance on transmission levels.

## 2 Model description and parameterisation

We adapted the stochastic formulation of the population-based gHAT transmission and control model described in [9], which builds on previous work [10, 11]. A schematic of the model is shown in Figure 1, and a description of the corresponding state variables is given in Table 1. The model is based on a system of ordinary differential equations that include tsetse flies, humans in multiple disease stages, and two risk settings. The low risk and high risk settings represent the ‘village’ and ‘plantation’ settings, respectively. Some individuals are modelled to travel between the two, and we assume those in the high risk setting do not participate in active screening, as there is an opportunity cost to screening [12, 13]. Animals are considered to receive bites but are assumed to not contribute to parasite transmission. A deterministic version of the model was fitted using a Bayesian approach to screening data and staged reported case data from both active and passive surveillance from Mushie territory in the DRC from 2000 to 2018 [14, 15]. This was then projected forward using stochastic simulations to 2040, with fexinidazole being introduced in 2021. Projections used the mean number of people screened annually by active screening between 2014 and 2018, leading to a decreasing active screening rate as the population size increases. The passive detection rate, i.e. the rate at which patients are removed from the infected compartments due to seeking treatment at a health centre, was kept constant at the 2018 rate from 2019 onwards. Parameters such as the ratio of humans in the high to low risk settings, the rate of passive detection between 2000 and 2018, the ratio of vectors to humans, and the diagnostic specificity were fitted using were fitted using a Bayesian approach and an adaptive Metropolis-Hastings Markov chain Monte Carlo approach was used to sample from the posterior distributions. Forward simulations were then run from 2000 to 2040 using the direct method of the Gillespie algorithm [16], implemented in a combination of R and C++ using the ‘Rcpp’ package. Further information on model assumptions, paramerisation and calibration can be found in the supplementary material.

**Figure 1:**
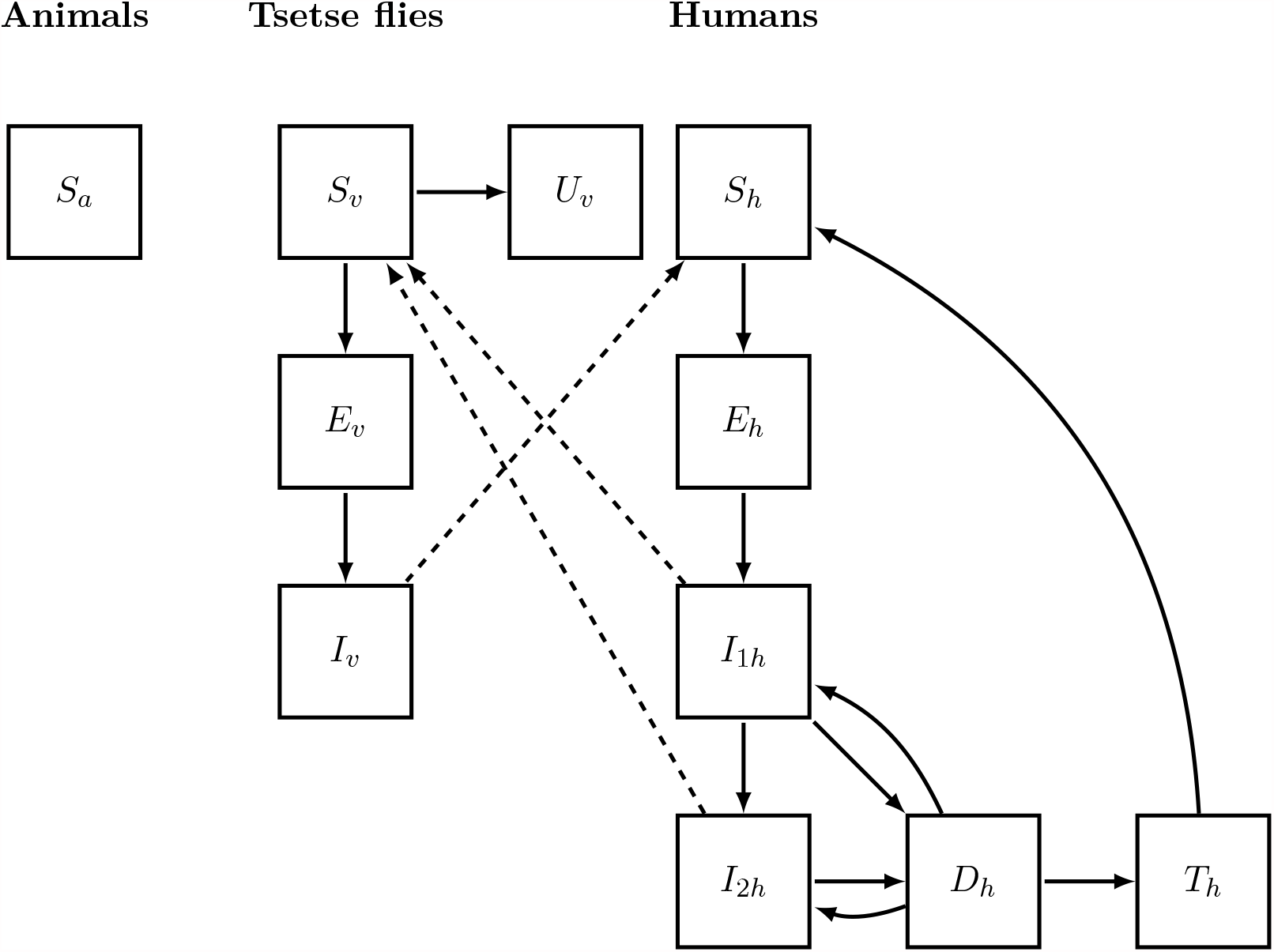
Overview of the model compartments. Solid lines depict transitions between compartments, while dashed lines represent transmission. State variable descriptions can be found in Table 1. Animals can receive bites from tsetse flies, but are assumed to not carry or transmit the disease.

**Table 1:** Description of model state variables. These variables exist for both the high and low risk settings.

| Variable | Description |
| --- | --- |
| $S_a$ | Number of animals |
| $S_v$ | Number of susceptible vectors |
| $E_v$ | Number of exposed vectors |
| $I_v$ | Number of infected vectors |
| $U_v$ | Number of non-teneral vectors |
| $S_h$ | Number of susceptible humans |
| $E_h$ | Number of exposed humans |
| $I_{1h}$ | Number of infected humans in stage 1 |
| $I_{2h}$ | Number of infected humans in stage 2 |
| $D_h$ | Number of diagnosed humans |
| $T_h$ | Number of treated humans |

## 3 Fexinidazole parameters and assumptions

Currently, it is unclear what proportion of patients in each stage will receive fexinidazole versus pentamidine or NECT, and the likely level of compliance. Thus, we have considered a range of values for the three parameters used to model fexinidazole treatment:

- **Compliance:** The proportion of patients receiving fexinidazole who comply with treatment guidelines sufficiently to be treated. The values simulated here were 25%, 50%, 75% and 100%.
- **Stage 1 access:** The proportion of patients in stage 1 of the disease, detected either through active screening or passive surveillance, receiving fexinidazole rather than the current treatment. The values simulated here were 25%, 50%, 75% and 100%.
- **Stage 2 access:** The proportion of patients in stage 2 of the disease, detected either through active screening or passive surveillance, receiving fexinidazole rather than the current treatment. The values simulated here were 25%, 50%, and 75%.

Efficacy of fexinidazole and current treatmentare both assumed to be 100%, though in a head-to-head comparative study between fexinidazole and NECT for stage 2 HAT, NECT was found to be 98% effective after 18 months, and fexinidazole 91% effective [17]. In this model, any reduction in efficacy would have the same impact as a reduction in compliance, as compliance captures the probability that someone who is diagnosed is completely cured of the disease.

Five scenarios were selected from the full-factorial combination described above for further analysis (Table 2). In the model, a 100% compliance to fexinidazole has the same effect on transmission as the recommended treatment that is currently in use, independent of the proportions of stage 1 or stage 2 cases treated with fexinidazole; this is the so called “full compliance” scenario in Table 2. “Worst case” scenarios were scenarios with the lowest compliance (25%) and either widespread use of fexinidazole for stage 1 cases (100%), for stage 2 cases (75%), or both. An additional scenario with widespread access to fexinidazole in both stages (75% for stage 1 and 50% for stage 2) and high compliance (75%) was also included as a baseline for what may be the usage of the drug given WHO guidelines are followed (“high compliance” scenario). Widespread access was considered appropriate as active detection leads to cases being reported earlier in the disease progression. This is reflected in the high ratio of cases detected through active screening versus passive surveillance seen in the data. Thus, we expect the majority of diagnosed cases will be detected before late stage 2, and so can be treated with fexinidazole.

**Table 2:** Five scenarios were considered in modelling the impact of fexinidazole use on the transmission of gHAT.

| Scenario | Compliance | Access in stage 1 | Access in stage 2 | Description |
| --- | --- | --- | --- | --- |
| Full compliance | 100% | 75% | 50% | Perfect compliance, same as current treatment |
| High compliance | 75% | 75% | 50% | Imperfect, but high, compliance, with widespread access to fexinidazole |
| Worst case — stage 1 | 25% | 100% | 25% | Poor compliance and widespread use for stage 1 patients |
| Worst case — stage 2 | 25% | 25% | 75% | Poor compliance and widespread use for stage 2 patients |
| Worst case — both stages | 25% | 100% | 75% | Poor compliance and widespread use for patients in both stages |

It is possible that with a logistically simpler and less unpleasant treatment, a larger number of health facilities will be able to administer the treatment. Additionally, the possibility to avoid a lumbar puncture will likely lead to less stigma around being tested for the disease. This may lead to an increase in the rate of passive detection. Increases of 20%, 50% and 100% in the passive detection rate in both stages from 2021 onward were explored. The corresponding mean increases in the proportion of patients accessing treatment in 2021 are given in Table 3. Note that these numbers are not identical since patients can be removed from each compartment due to other processes such as active screening (low-risk setting only), disease progression and death. These values also change over time in the low risk setting as the active surveillance rate decreases over time.

**Table 3:** Increases in passive detection rate and corresponding mean increase in the proportion of patients receiving treatment in each setting and disease stage.

| Setting | Increase in passive detection rate (%) | Corresponding increase in patients of that setting and stage receiving treatment (%) |
| --- | --- | --- |
| Low risk - stage 1 | [20, 50, 100] | [16, 37, 68] |
| Low risk - stage 2 | [20, 50, 100] | [8, 20, 33] |
| High risk - stage 1 | [20, 50, 100] | [14, 33, 60] |
| High risk - stage 2 | [20, 50, 100] | [17, 40, 76] |

## 4 Results

### 4.1 Impact of reduced compliance

As expected, the greatest reduction in incidence between 2021 and 2040 is achieved with perfect compliance and is at least equivalent to continuing the use of the current treatment regimen (Fig. 2). In comparison, in the worst case scenario (low compliance and extended use of fexinidazole in both stages), we see a delay of 7 years in achieving elimination of transmission when considering the median incidence, as compared to the full compliance scenario. Widespread use of fexinidazole in stage 1 in low compliance scenarios (worst case - stage 1 and worst case - both stages) has a larger overall negative impact on transmission, likely because stage 1 cases that are not effectively treated can potentially transmit the disease to susceptible individuals for a longer period than stage 2 non-compliants. While attention should be paid towards treatment adherence regardless of stage in line with WHO recommendations [4], this result suggests that compliance in stage 1 patients is especially important for reducing further transmission of the disease.

**Figure 2:**
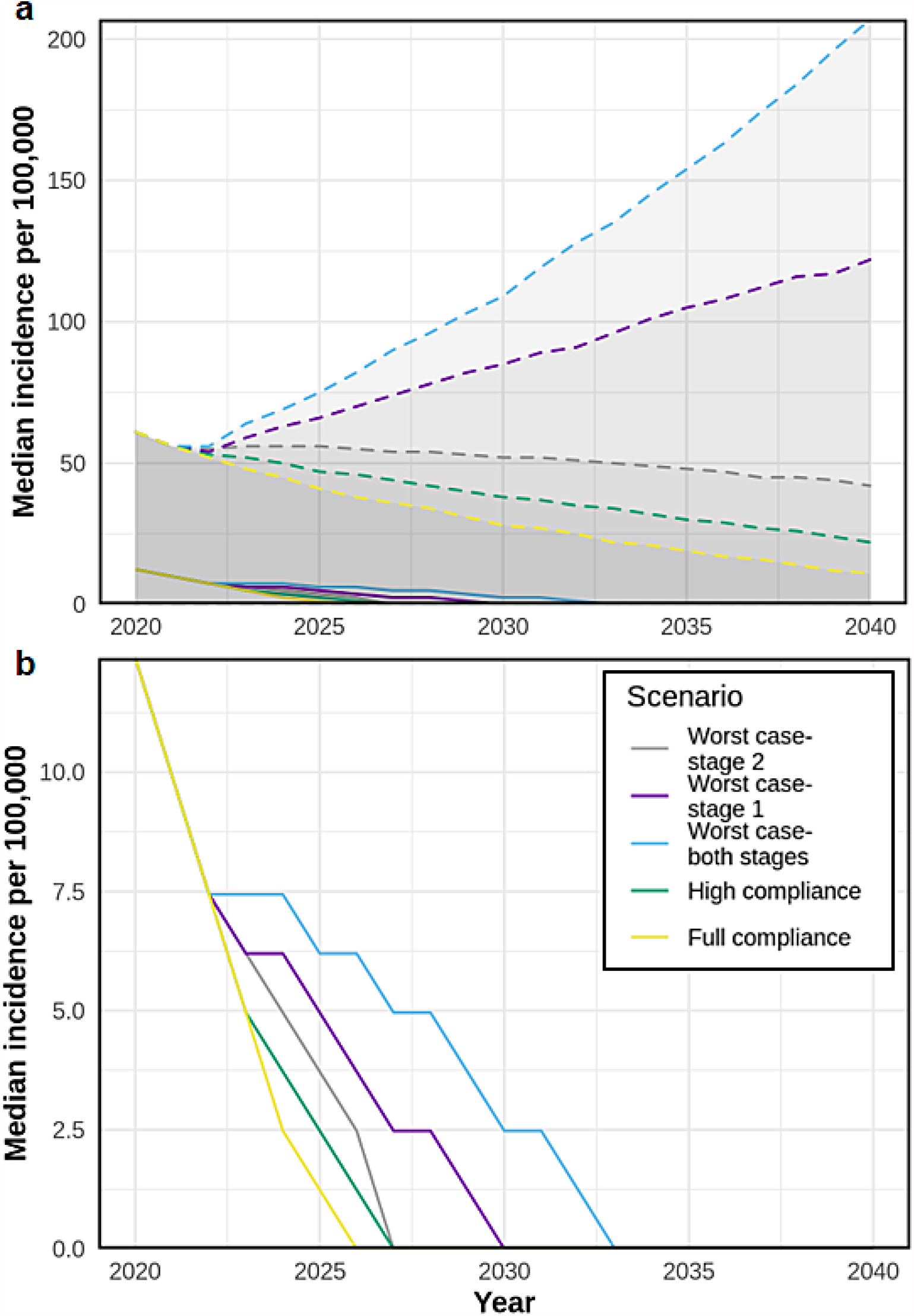
gHAT incidence per 100,000 at various fexinidazole compliance and use levels. Fexinidazole has been modelled to be introduced from 2021. Descriptions of the scenarios can be found in Table 2. **(a)** The median incidence including the 95% confidence intervals (shaded area bounded by dashed lines of the same colour). **(b)** The median incidence per 100,000 for each scenario. Note the different scales on the y-axis for the two plots.

While the median simulated incidence is declining for all scenarios (Fig. 2b), there are some potential parameter sets where low compliance leads to an increase in cases (Fig 2a). This is because the observed historic data is compatible with parameters that would lead to an increase in disease incidence if compliance with treatment was low. This result suggests that the situation should be monitored closely after the introduction of fexinidazole, and an increase in cases may indicate that drug compliance is low.

### 4.2 Improvements in passive detection rate

The issue of non-compliance is likely to be countered by the increased number of patients who access treatment, particularly those in an early stage of the disease. Increasing the passive detection rate by ∼20% is expected to be sufficient to ensure a similar trend in incidence and probability of elimination of transmission in the “high compliance” scenario as expected with the current treatment (Fig. 3, black dashed line versus green line).

**Figure 3:**
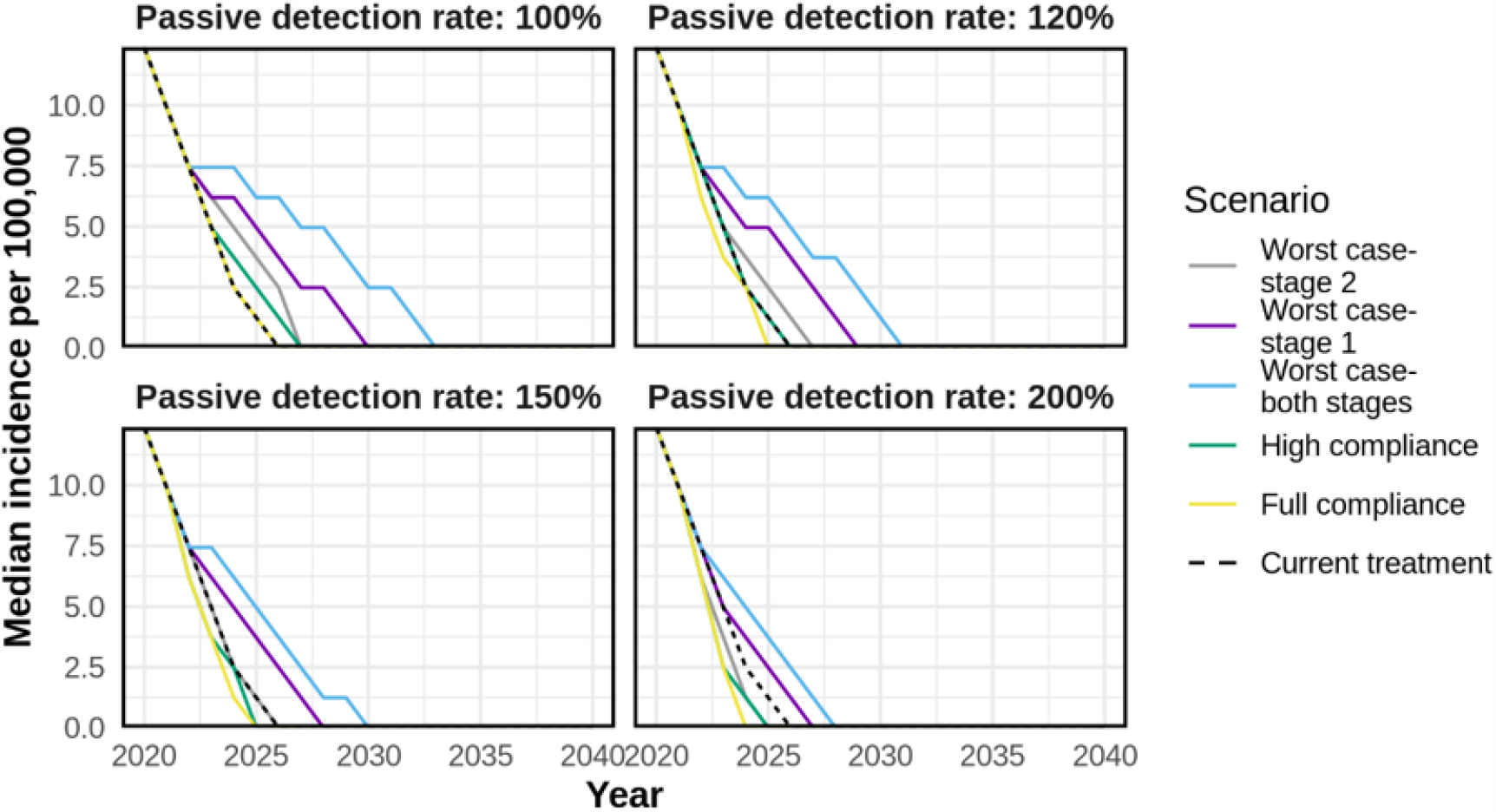
The time-series of median incidence per 100,000 population at increasing passive detection rates. Descriptions of the scenarios can be found in Table 2. The dashed black line corresponds to the current treatment (equivalent to 100% compliance and no increase in the passive detection rate). Fexinidazole has been modelled to be introduced from 2021.

Nonetheless, if compliance is low and access to fexinidazole for both stages is high, even after doubling the passive detection rate, a substantial drop in the probability of the elimination of transmission over time is expected. The probability of elimination of transmission (EOT) for any given year is defined as the proportion of simulations which have reached zero exposed and infected humans and vectors by that year. The probability of EOT by 2030 is 65% if the treatment regimen that is currently in use is continued with no increase in passive detection (Fig. 4, black dashed line). However, in the worst case scenario, even when the passive detection rate is 200% of the current passive detection rate, the WHO target of EOT by 2030 is achieved in only 50% of the simulations (Fig. 4, light blue line).

**Figure 4:**
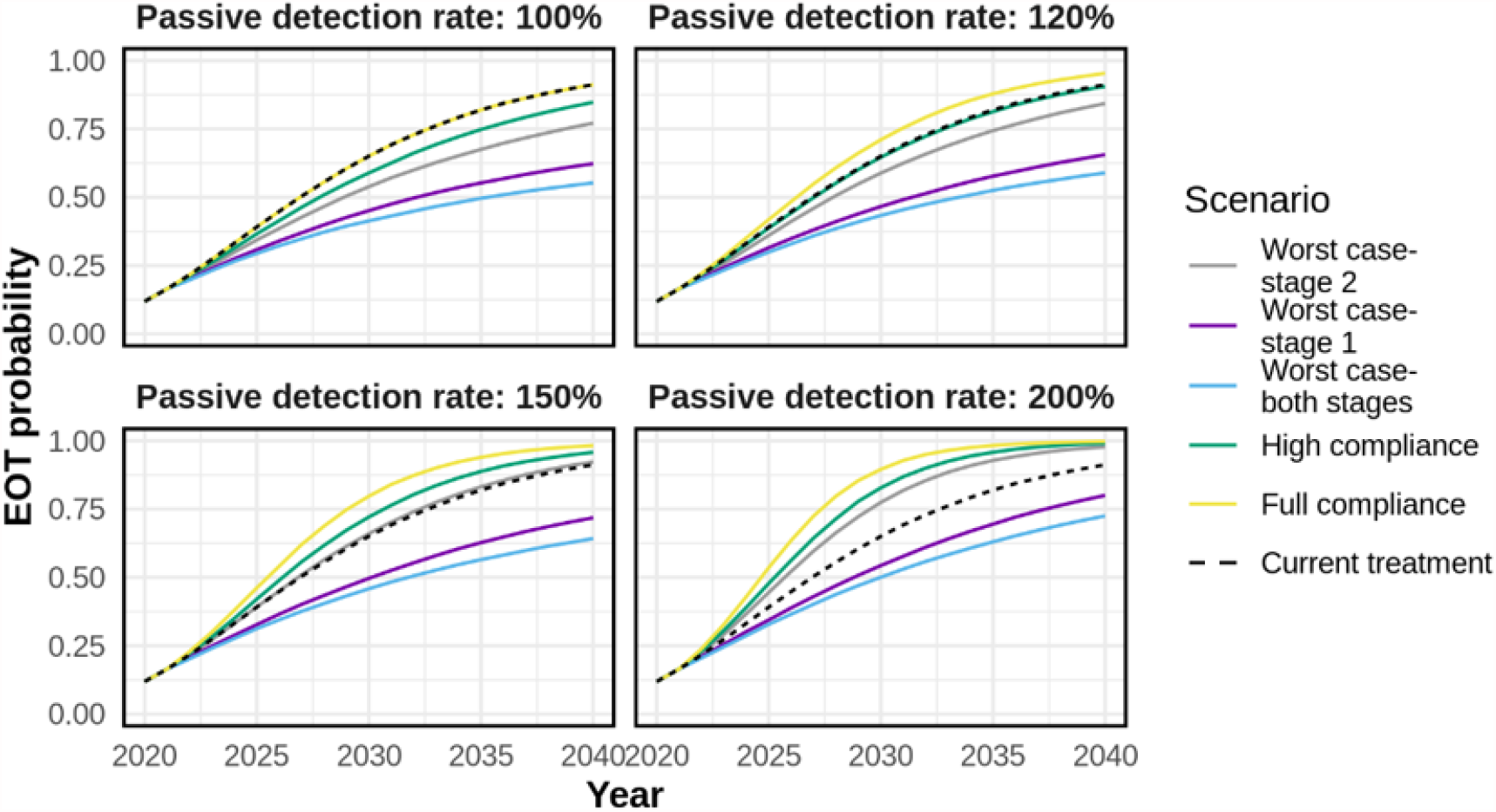
The probability of the elimination of transmission (EOT) at increasing passive detection rates. This probability of EOT reflects our uncertainty in the setting (the uncertainty in parameter values from our best fits to the data) and inherent stochastic variation. Mathematically, it is calculated as the proportion of all simulation runs (including different parameterisations and random seeds) that have reached zero exposed and infected humans and vectors by that year. Descriptions of the scenarios can be found in Table 2. The dashed black line corresponds to the current treatment (equivalent to 100% compliance and no increase in the passive detection rate). Fexinidazole has been modelled to be introduced from 2021.

## 5 Conclusions and discussion

The effect of fexinidazole on gHAT transmission depends on treatment adherence and the proportion of diagnosed patients that receive the drug. If compliance is low, particularly in stage 1 patients, fexinidazole could have a substantial negative impact on the decline in gHAT incidence seen in recent years. The possibility of achieving the WHO 2030 goal of elimination of transmission is expected to decrease with low compliance and widespread use of fexinidazole. This would be due to the higher number of incompletely treated patients potentially contributing to transmission, with stage 1 patients typically contributing for longer to further transmission than stage 2 patients. However, if compliance is high, especially if it also leads to more patients arriving at health facilities for diagnosis, then the impact of fexinidazole may be a positive one. In a near-elimination disease setting such as the case of HAT in the DRC, it is expected that at some point, active screening is likely to be scaled back as it is a resource-intensive intervention. With this, the relative importance of passive detection will increase, as that will be the main mechanism by which cases are detected and treated. An oral treatment that can be administered in the primary care setting, such as fexinidazole, may remove some barriers to self-presentation at health centres. Firstly, patients would have to travel less far on average to receive treatment if more health facilities could provide HAT treatment, particularly for the second stage of the disease [18]. In 2018, it was estimated that while 696 facilities in the DRC could provide a diagnosis of HAT, only 191 health facilities could provide treatment for second-stage HAT with NECT [14]. Secondly, fear of lumbar punctures was also identified as a reason why some patients avoid HAT screening [12]. This would no longer be necessary for non-severe HAT cases that qualify for fexinidazole treatment. Other factors that may improve passive detection include ensuring that indirect costs for treatment are affordable [12, 19]. Removing the need for a lumbar puncture and hospitalisation would help to reduce the cost faced by patients for HAT treatment. Finally, administering fexinidazole would require fewer healthcare resources than the current treatment pathway, which has been previously highlighted as a challenge to controlling the disease [20].

WHO guidelines recommend fexinidazole only when there is confidence in concomitant food intake and confidence in full adherence. Following these guidelines would likely lead to high levels of compliance, averting the worst case scenarios presented in this study. Until now, no study has reported on adherence to fexinidazole treatment, and studies on adherence to other oral treatment in rural African settings for diseases such as malaria and HIV treatment show a high variability in compliance, which depends on multiple factors including gender, age, education level and side effects among others [21, 22, 23, 24]. A systematic review of interventions to promote patient adherence to antimalarial medication found a significant increase in adherence when treatment was observed by a medical professional [25]. It is difficult to predict at this stage the likely impact of an oral treatment such as fexinidazole on the passive detection rate. However, if it increases the rate at which patients seek diagnosis and treatment, then the effects of lower treatment compliance may be mitigated by increases in the proportion of patients receiving treatment.

It is worth noting that non-compliance will lead to patients being reported as treated when they may in fact still be infectious. Thus, the number of successfully treated cases may differ from the number of cases reported through official channels. In line with WHO guidelines, we would recommend follow up of cases treated by fexinidazole to confirm that the treatment was successful and to detect any relapses early. However, this may prove challenging in rural settings where gHAT is prevalent.

One limitation of this study is that it does not consider disruption to ongoing control activities due to the 2019 coronavirus disease (COVID-19) pandemic. Active screening was suspended in the DRC in 2020 due to the pandemic. A previous study conducted with this model and another stochastic model that was independently developed for gHAT suggested that if the disruption was to continue until the end of 2021, a delay of 2-3 years in achieving EOT would be expected [26]. The likely effect of this disruption would be to reduce the probabilities of EOT by 2030 presented in this paper, but the overall trends would remain the same.

In conclusion, this study highlights the need for careful monitoring of compliance with the use of an oral medication such as fexinidazole for the treatment of HAT. Reduced compliance is expected to lead to delays in achieving HAT elimination, but if the case detection rate increases, this may be sufficient to offset any increases in transmission due to poor treatment adherence. Further studies are required, particularly in the field setting, to better quantify the likely effect of fexinidazole on drug compliance and the rate of passive detection.

## Data Availability

The data underlying the results presented in the study are available upon request, to researchers who meet the required criteria, from the World Health Organization HAT Atlas (contact via Dr. Jose Ramon Franco Minguell -).

