## Supplementary information for "Modelling the impact of fexinidazole use on human African trypanosomiasis transmission in the Democratic Republic of Congo"

### 1 Model description

#### 1.1 Deterministic model

This description is adapted from [1, Supplementary data].

The deterministic model used here was presented and described in [2] and is a variant of the HAT transmission model originally published in [3] and [4]. The model consists of a system of coupled ordinary differential equations (ODEs), with compartments for tsetse, animal and human populations. These three different host types are modelled for two different settings corresponding to a low transmission area (e.g. the village, $L$) and a high transmission area (such as river banks or plantations, $H$) that enable accounting for heterogeneity in exposure to tsetse bites. The population size for tsetse, animal or humans in each setting $i$ ($i = \{L, H\}$) is assumed to be stable by allowing the associated birth terms to compensate deaths in all the compartments. Tsetse and animal populations always stay within their setting (for example, tsetse in low transmission settings always remain in the low transmission setting and animals in high transmission settings always remain in the high transmission setting). Similarly, humans in low transmission settings always remain in low transmission setting. However, humans in the high transmission setting move back and forth between the high and low transmission settings spending a fixed amount of time in each one (to model, for example, the movement of high risk individuals between villages and plantations) — as shown in Figure 1.

Five compartments describe humans in any of the two settings: susceptible ($S_{hi}$); exposed or incubating ($E_{hi}$); infected with the first stage of the disease ($I_{h1i}$); infected with the second stage of the disease, where trypanosomes have reached the cerebro-spinal fluid ($I_{h2i}$); and treated ($T_{hi}$). The total human population in setting $i$ is $N_{hi} = S_{hi} + E_{hi} + I_{h1i} + I_{h2i} + T_{hi}$. Humans can simultaneously belong in the diagnosed compartment ($D_{hi}$)

and one of the infected stages, from which, depending on drug compliance, they may either move on to the treated compartment, or remain in the infected compartment.

Tsetse populations are divided into susceptible ($S_{vi}$); teneral ($U_{vi}$); exposed ($E_{vi}$); and infected ($I_{vi}$), so that the vector population is $N_{vi} = S_{vi} + U_{vi} + E_{vi} + I_{vi}$.

As in [1], in this model implementation: *i)* animals do not contribute to transmission, thus animal populations are modelled as constant parameters, $N_{ai}$, and only form a sink for tsetse bite; *ii)* both stages (rather than only stage 1) of the disease are exposed to tsetse fly bites; *iii)* an additional compartment in the vector dynamics, $U_i$, accounts for the teneral effect — a reduction of infectivity with time — such that on average tsetse are only infectious for the first five days after emergence. These changes were made with respect to the original version [3] to provide a more realistic representation of the transmission dynamics. A schematic of the model is shown in Figure 1.

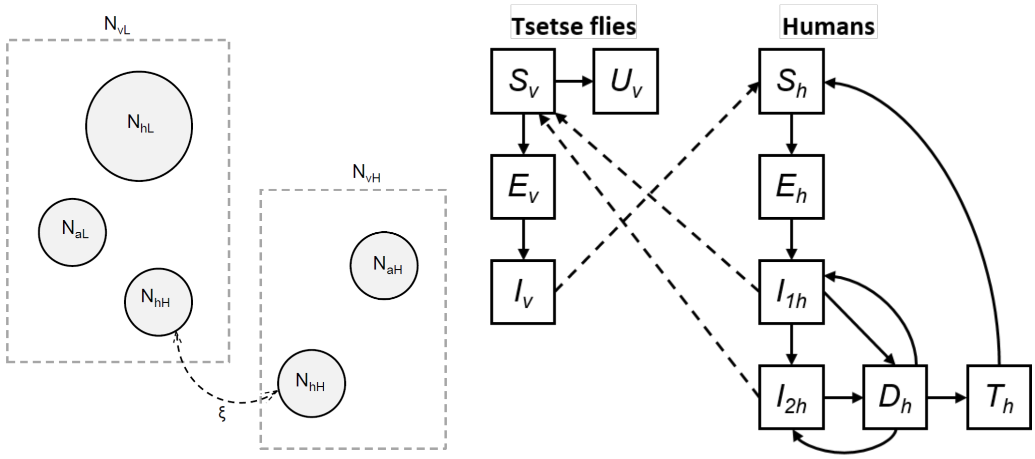


**Figure 1: Schematic of the model.** Left: model population structure. Human populations are composed by a stationary population ($N_{hL}$) that remains in low exposure habitats (e.g., a village), and a smaller population ($N_{hH}$) which commute and spend a proportion $\xi$ of their time in a potentially high exposure setting (e.g., a plantation). Each habitat also contains tsetse ($N_{vL}$ and $N_{vH}$) and non-human vertebrate animal populations ($N_{aL}$ and $N_{aH}$). Right: schematic of infection dynamics, subscripts $i = \{L, H\}$ were removed for easy reading. Compartmental diagram highlights the transmissions between states of infection of the tsetse and human populations, with solid lines indicating transition between compartments, and dashed lines representing transmission rates. Animals cannot transmit infection thus acting as a sink for tsetse bite. Note that in the low-risk transmission setting, both human populations are exposed to tsetse bites. Figure adapted from [3].

##### 1.1.1 Stochastic implementation

This description is adapted from [1, Supplementary data].

Epidemiological deterministic models, including previous implementations of this model [3, 5, 4], have the shortcoming that they do not capture rare events and do no account for the discrete nature of populations, and are therefore unable to reproduce the transition between extremely low prevalence and zero transmission. For such situations, a discrete stochastic model formulation is more suitable as it captures the stochastic nature of events involved in transmission dynamics while producing integer outputs (e.g. number of cases and new infections here) that enable a clearer definition of elimination of transmission (and subsequent forecasting of elimination timelines) than in deterministic ODE models where arbitrary thresholds must be defined.

In the stochastic formulation of the ODE model described in 1.1, we model all human host, animal host and tsetse fly populations as discrete numbers, and all individuals move probabilistically between compartments at varying intervals of time. Any process governing the HAT transmission dynamics is considered stochastic, with terms in the compartmental model being now considered as probabilities at which an event occurs.

We implemented the direct method of the stochastic simulation algorithm (SSA; also known as Gillespie method [6]). In the direct method of the SSA, all possible events in the HAT transmission dynamics have an associated rate given by the associated term in the deterministic ODEs. For example, if $\gamma$ represents the rate at which humans infected in the stage 1 of the disease ($I_1$) move to the second stage of the disease ($I_2$), thus $\gamma I_1$ represents the rate $R$ for the event "progression to stage 2 of the disease".

In order to simulate one stochastic realisation under the direct method, for $i$ possible events with associated rate $R_i$, at any time $t$:
($a$) we determine the time $t + \tau$ at which the next event happens, with $\tau$ an exponentially distributed random number scaled by the sum of all process rates, $\Sigma_i R_i$; and
($b$) we decide which that event will be: the event that happens next is obtained through drawing a process randomly from all possible processes according to their respective probabilities given by $R_i / \sum_j R_j$.

In the present analysis, 1000 posterior parameter sets (see section 2) were used along with the fixed parameters to obtain 100,000 realisations of the stochastic model (100 realisations for each parameter set).

Simulations up to 2000 were deterministic, i.e. by numerical solving the ODE system, with further projections using the stochastic implementation described in this section.

The deterministic model output at 2000 was scaled up to follow the population growth trajectory of Mushie territory and rounded to integer values before running stochastic simulations of the forward discrete model. Simulations from 2000 to 2018 were run using parameters calibrated to data, and then projected forward until 2040 including reductions in the active screening rate (see section 1.2) and maintaining the passive detection rate seen in 2018.

The discrete stochastic model was run until 2040 in order to allow evaluation of elimination of transmission (EOT) (Figure 4 in the main text), with EOT defined as the point where there are no exposed or infected humans or vectors present in the simulation.

#### 1.2 Screening

This description is adapted from [2, Supplementary data].

Active screening was modelled via a constant annual active detection rate $r_{as}$ that removes infected people only from the low risk setting. As in previous works, we followed [7] to relate a proportion, $d$, of humans effectively screened in a given year and the annual removal rate $r_{as}$ as $d = 1 - e^{-r_{as}}$, leading to $r_{as} = -\ln(1 - d)$. For the model fitting, screening levels were informed from data, and estimates for the health zone population in 2015 were taken from [8], and projected backwards and forward in time assuming a 3% annual growth rate. For model projections, the mean number of people screened from the last 5 years of available data (2014-2018) was used to define $d$, with an ongoing 3% growth rate in the total population (leading to a continuing decrease in the proportion of the population screened).

Passive detection is represented by a continuous stage-specific detection rate, $r1$ and $r2$ for stage 1 and stage 2 respectively, and removes infected people from both low- and high-risk settings. Relying on previous work [4], improvement to passive detection was assumed for the data period. We modelled improvement for the number of years $y > 0$ after 2000 as a logistic function:

$$r1(y) = 365 \times r1_{\text{const}} + \frac{\Delta r_1}{1 + \exp(-\alpha_{\text{pd}}(y - x_0 + 1))},$$

$$r2(y) = 365 \times r1_{\text{const}} \times c2 + \frac{\Delta r_1 + \Delta r_2}{1 + \exp(-\alpha_{\text{pd}}(y - x_0 + 1))},$$

where $r1_{\text{const}}$ and $c_2 \times r1_{\text{const}}$ are the constant daily passive detection rates in stage 1 and stage 2 respectively for any time before 2000, and $\Delta r_1$, $\Delta r_2$, $\alpha_{\text{pd}}$ and $x_0$ are parameters defining the profile of the logistic curve. All parameters in the expression above are fitted to the health zone level data. For model projections, we assumed passive detection rates $r1$ and $r2$ continue at the highest level from 2000-2018.

Additionally, due to changes to the method of confirming positive HAT cases, including video evidence of moving parasites, a perfect specificity of 100% was assumed from 2017 onwards. This is reflected in the cases seen, as there is a drop in reported cases observed from 2017 onwards.

Our model assumes that before 2000 only passive detection was ongoing, at constant rates, and that active screening activities started in 2000, the initial year for which there is available data on active screening.

### 2 Fitting procedure

Twelve parameters were fitted using annual case data from Mushie territory for the period 2000–2018. The deterministic model was first run to reach equilibrium prevalence of infection assuming only constant passive screening before 2000, when the fitting starts. The data that was fitted separated reported cases into those from active screening and passive detection, and provided information on staging in the years from 2015 onward. Fitting was performed via an adaptive Metropolis-Hastings Markov chain Monte Carlo (MCMC) approach using the following log-likelihood function:

$$
\begin{aligned}
LL(\theta|x) = & \log(P(x|\theta)) \\
\propto & \sum_{i=2000}^{2018} \Bigg( \log\left[\text{NegBin}\left(A_{d1}(i) + A_{d2}(i); A_{m1}(i) + A_{m2}(i), \kappa_{\text{AS}}\right)\right] \\
& + \log\left[\text{NegBin}\left(P_{d1}(i) + P_{d2}(i); P_{m1}(i) + P_{m2}(i), \kappa_{\text{PD}}\right)\right] \Bigg) \\
& + \sum_{i=2015}^{2018} \Bigg( \log\left[\text{Bin}\left(P_{d1}(i); P_{d1}(i) + P_{d2}(i), \frac{P_{m1}(i)}{P_{m1}(i) + P_{m2}(i)}\right)\right] \\
& + \log\left[\text{Bin}\left(A_{d1}(i); A_{d1}(i) + A_{d2}(i), \frac{A_{m1}(i)}{A_{m1}(i) + A_{m2}(i)}\right)\right] \Bigg),
\end{aligned}
$$

where $A_{d1}$: stage 1 reported cases (active screening); $A_{d2}$: stage 2 reported cases (active screening); $P_{d1}$: stage 1 reported cases (passive detection); $P_{d2}$: stage 2 reported cases (passive detection); $A_{m1}$: stage 1 reported cases from the model (active screening); $A_{m2}$: stage 2 reported cases from the model (active screening); $P_{m1}$: stage 1 reported cases from the model (passive surveillance); $P_{m2}$: stage 2 reported cases from the model (passive surveillance); $\kappa_{AS}$: shape parameter for the negative binomial distribution for annual number of cases detected through active screening; and $\kappa_{PD}$: shape parameter for the negative binomial distribution for annual number of cases detected through passive detection.

The two terms in the first sum represent the total number of active and passive cases in the data, respectively, modeled as a negative binomial with mean equal to the number of cases from the differential equation model. The two terms in the second sum represent the proportion of cases in stage 1, modeled as a binomial in which the probability of stage 1 is the proportion from the differential equation model (and the number of trials is from the data of total number of cases). Since we only have staged data from 2015 onwards, those terms only contribute to the log likelihood in those years.

To sample from the posterior distribution, determined by the likelihood function and the prior distributions, we used an adaptive Metropolis-Hastings MCMC algorithm – the accelerated shaping algorithm (unpublished, Spencer). We ran two independent chains of the algorithm to corroborate convergence for the sampling. We used a burn-in period of 2000 steps and then ran the chain for 20,000 steps, which was thinned to every other sample. For the proposal distribution, we used a multivariate Normal distribution (truncated with the bounds given in Table 1), with a covariance matrix that adapts to predict the shape and scale of the posterior distribution as the algorithm proceeds. The adaptation improves the efficiency of proposing new samples so that there are neither excessive rejections nor acceptances in the algorithm. Finally, to improve mixing further, we used the two covariance matrices from this first set of runs in a second round of two independent chains with the same burn-in and sampling strategy. With this second set, we visually checked that there was good mixing. The parameters used in the forward projections are based on the first chain of this second set of runs.

#### 2.1 Fixed parameters, priors and posterior distributions

Descriptions and values of all fixed parameters are given in Table 2, and descriptions and prior distributions for all fitted parameters are given in Table 1 and Table 2 respectively.

| **Parameter** | **Unit** | **Prior Distribution and Bounds** |
|---|---|---|
| $\kappa$ | - | Unif$[0, 1]$ |
| log(VHL) | - | N$(1.1, 0.05)$ in [0,log(100)] |
| log($c_1$) | - | Gamma$(1, 1)$ in [0,log(50)] |
| logit(spec) | - | Unif[logit(0.998), logit(0.9999)] |
| $r1_{\text{const}}$ | day$^{-1}$ | Unif$[0, 10^{-3}]$ |
| log($c_2$) | - | Gamma$(1, 1)$ in [0,log(50)] |
| $\Delta r_1$ | year$^{-1}$ | Unif$[0, 2.5]$ |
| $\Delta r_2$ | year$^{-1}$ | Unif$[0, 2.5]$ |
| $x_0$ | - | Gamma$(10, 0.06)$ in $[0, 19]$ |
| $\alpha_{\text{pd}}$ | - | Unif$[0.1, 5]$ |
| $\kappa_{\text{as}}$ | - | Gamma$(23.5, 3)$ |
| $\kappa_{\text{pd}}$ | - | Gamma$(23.5, 3)$ |

**Table 1: Priors for fitted parameters.** Non-uniform priors were additionally truncated with values given in brackets. Gamma priors are written with arguments of shape and scale.

**Table 2: Model parameterisation (posteriors of fitted parameters).** Notation, a brief description, and representative percentiles of the posterior distributions for fitted parameters. Here logarithm always refers to the natural logarithm.

| Notation | Description | Posterior (median [95% CI]) |
|---|---|---|
| $\kappa$ | Ratio of humans in the high- to low-exposure environment | 8.25 [ 3.82,11.6 ]$\times 10^{-2}$ |
| log(VHL) | Log ratio of vectors to humans in low-exposure environment (VHL) | 1.053 [0.974, 1.445] |
| log($c_1$) | Log ratio of the ratio of vectors to humans in the high exposure environment to the ratio of vectors to humans in the low exposure environment | 6.585 [0.547, 17.02]$\times 10^{-2}$ |
| spec* | Diagnostic specificity (active screening) | 0.9991 [0.9990, 0.9992] |
| $r1_{\text{const}}$ | Daily passive detection rate for stage 1 (pre-2000) | 3.62 [1.92,5.78]$\times 10^{-4}$ |
| log($c_2$) | Log ratio of passive detection for stage 2 to stage 1 (pre-2000) | 0.559 [0.036, 1.773] |
| $\Delta r_1$ | Amount passive detection in stage 1 improves | 9.978 [4.402, 17.16]$\times 10^{-2}$ |
| $\Delta r_2$ | Amount passive detection in stage 2 improves (in addition to improvement of stage 1) | 1.444 [0.646, 2.314] |
| $x_0$ | Turning point (years since 1999) for logistic improvement in passive detection | 13.24 [6.44, 18.76] |
| $\alpha_{\text{pd}}$ | Steepness in logistic improvement of passive detection | 0.578 [0.182, 0.962] |
| $\kappa_{\text{as}}$ | Overdispersion parameter (active screening) | 34.04 [19.56, 56.49] |
| $\kappa_{\text{pd}}$ | Overdispersion parameter (passive detection) | 61.45 [39.13, 91.33] |

* The specificity parameter, spec, was sampled in the logit scale, however posterior estimates are shown here in the model scale for clarity.

#### 2.2 MCMC outputs

The MCMC outputs shown in Figures 2 correspond to 10,000 post burn-in samples.

##### 2.2.1 Posterior densities

.

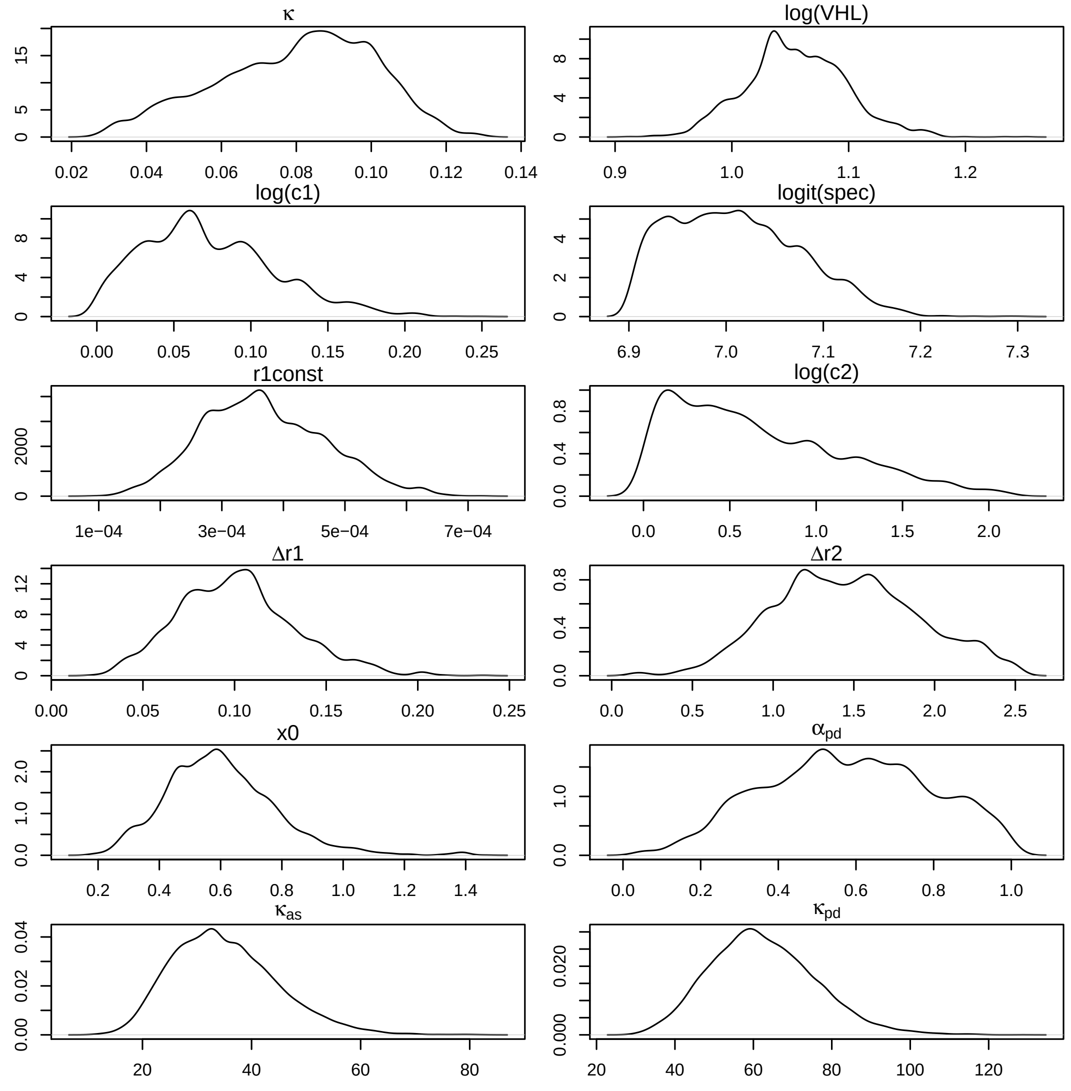


**Figure 2:** Posterior density of fitted parameters for Mushie territory.

##### 2.2.2 Model fit to reported case data

Figure 3 shows the model fit to Mushie territory. The deterministic model simulations used 10,000 MCMC posterior samples.

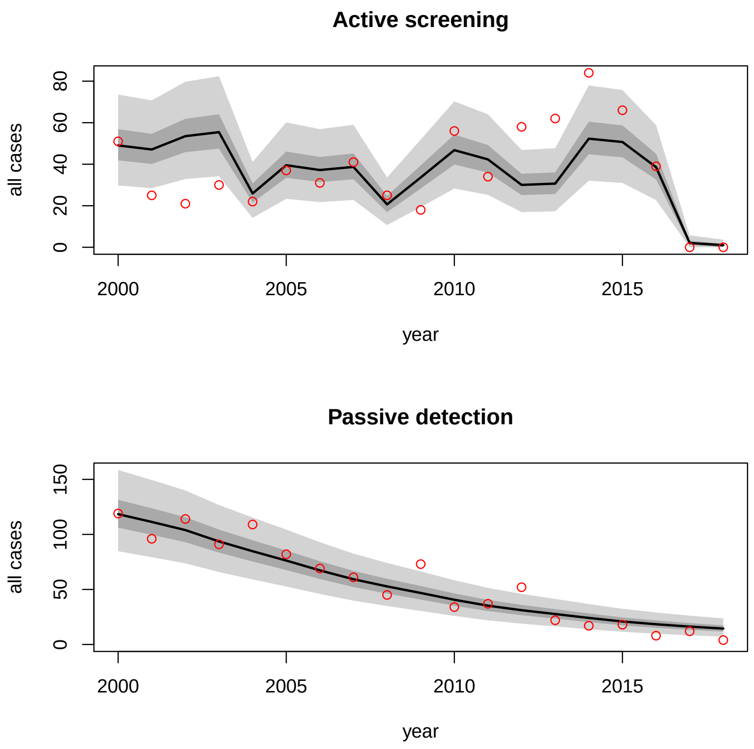


**Figure 3:** Model fit to reported case data for Mushie territory. Shaded regions indicate (2.5,97.5) and (25,75) percentiles.

[8] OCHA Office for the Coordination of Humanitarian Affairs. *Journées Nationales de Vaccination (JNV) Activités de vaccination supplémentaire , RDC*, Accessed May 2016.
